# Convalescent Plasma for COVID-19: A multicenter, randomized clinical trial

**DOI:** 10.1101/2020.08.26.20182444

**Authors:** C Avendaño-Solà, A Ramos-Martínez, E Muñez-Rubio, B Ruiz-Antorán, R Malo de Molina, F Torres, A Fernández-Cruz, A Callejas-Díaz, J Calderón, C Payares-Herrera, I Salcedo, I Romera, J Lora-Tamayo, M Mancheño-Losa, ML Paciello, C Villegas, V Estrada, I Saez-Serrano, ML Porras-Leal, MC Jarilla-Fernández, JR Paño-Pardo, JA Moreno-Chulilla, I Arrieta-Aldea, A Bosch, M Belhassen-Garcia, O López-Villar, A Ramos-Garrido, L Blanco, ME Madrigal-Sánchez, E Contreras, E Muñiz-Díaz, JM Domingo-Morera, I Casas-Flecha, M Pérez-Olmeda, Javier Garcia-Pérez, J Alcamí, JL Bueno, RF Duarte, for the ConPlas-19 Study Group

## Abstract

**Background:** Passive immunotherapy with convalescent plasma (CP) is a potential treatment for COVID-19 for which evidence from controlled clinical trials is lacking.

**Methods:** We conducted a multi-center, randomized clinical trial in patients hospitalized for COVID-19. All patients received standard of care treatment, including off-label use of marketed medicines, and were randomized 1:1 to receive one dose (250-300 mL) of CP from donors with IgG anti-SARS-CoV-2. The primary endpoint was the proportion of patients in categories 5, 6 or 7 of the COVID-19 ordinal scale at day 15.

**Results:** The trial was stopped after first interim analysis due to the fall in recruitment related to pandemic control. With 81 patients randomized, there were no patients progressing to mechanical ventilation or death among the 38 patients assigned to receive plasma (0%) versus 6 out of 43 patients (14%) progressing in control arm. Mortality rates were 0% vs 9.3% at days 15 and 29 for the active and control groups, respectively. No significant differences were found in secondary endpoints. At inclusion, patients had a median time of 8 days (IQR, 6-9) of symptoms and 49,4% of them were positive for anti-SARS-CoV-2 IgG antibodies.

**Conclusions:** Convalescent plasma could be superior to standard of care in avoiding progression to mechanical ventilation or death in hospitalized patients with COVID-19. The strong dependence of results on a limited number of events in the control group prevents drawing firm conclusions about CP efficacy from this trial. (Funded by *Instituto de Salud Carlos III;* NCT04345523).

## INTRODUCTION

Passive immunotherapy for COVID-19 with convalescent plasma (CP) is inexpensive, easily accessible, and has a rationale for use and experience in other viral epidemics. Overall, CP appears to be safe and potentially effective,^1^ although formal demonstration of its benefits in well controlled clinical trials is lacking. During the COVID-19 pandemic, the acute need for medical treatments has spread the use of unproven medicines based on the rationale for potential efficacy. Several antiviral agents have been empirically used (i.e lopinavir-ritonavir, hydroxychloroquine), to be subsequently withdrawn when they failed to demonstrate efficacy in randomized clinical trials (RCT).^2,3^ Remdesivir reduces the number of days of disease with no effect on hard outcomes such as progression to severe disease or death.^4^ In a large RCT,^5^ dexamethasone has shown a benefit in mortality in the subgroup of patients with oxygen support at the time of randomization and at least 7 days from symptoms onset. Regarding CP, a first RCT, prematurely stopped by logistic reasons, did not demonstrate benefit in COVID-19 patients with a median of 30 days of symptoms. However, a positive effect was suggested in the subgroup of non-critical patients.^6^ A second trial was also prematurely stopped after a first interim analysis showed that most patients already had high neutralizing antibody titers at the time of inclusion and that no statistical differences in outcomes were seen between treatment groups.^7^ The ConPlas-19 RCT was designed to demonstrate the efficacy and safety of CP used to prevent progression to severe disease or death in hospitalized patients with earlier forms of COVID-19.

## METHODS

### Study oversight

ConPlas-19 is a multicenter open-label RCT funded by the Spanish *Instituto de Salud Carlos III* (COV20/00072), conducted in 14 hospitals in Spain and registered at clinicaltrials.gov as NCT04345523. Enrollment of CP donors began on April 2, 2020. Patients enrollment began on April 4 and ended on July 10. All patients received standard of care (SOC), including all supportive and specific treatments with off-label marketed medicines used according to local or national recommendations, and were randomly assigned in a 1:1 ratio to receive a single unit of CP (250-300 mL).

CP donors complied with EU requirements for plasma donors^8^, had laboratory confirmed SARS-CoV-2 infection, anti-SARS-CoV-2 IgG (ratio ≥1.1 with the Euroimmun ELISA test; Euroimmun, Lübeck, Germany) and were asymptomatic for at least 14 days. A history of transfusion or pregnancy including miscarriages were exclusion criteria to minimize the risk of TRALI (Transfusion Related Acute Lung Injury). Neutralizing antibodies in CP were tested with a pseudovirus assay based on an HIV-defective platform expressing the spike protein from SARS-CoV-2 and by viral microneutralization test (VMNT) with infectious SARS-CoV-2 (references and methods in supplementary appendix). Neutralizing antibodies results were available at the end of the study and not used to select CP donors or units. Patients were eligible if hospitalized for laboratory-confirmed SARS-CoV-2 infection (RT-PCR) with either radiographic evidence of pulmonary infiltrates or clinical evidence plus SpO2 ≤94% on room air, and within 12 days from the onset of symptoms (fever or cough). Patients were excluded if already on mechanical ventilation (invasive or non-invasive) or high flow oxygen devices. A detailed description of inclusion and exclusion criteria as well as amendments to the conduct of the study are available in the supplementary appendix.

Oral informed consent (to avoid paper handling) was obtained from all the patients. Written witnessed consent was documented in the medical records and written consent by the patient was later obtained when feasible. The trial was approved by the Research Ethics Committee of the Hospital Universitario Puerta de Hierro Majadahonda and conducted in accordance with the principles of the Good Clinical Practice guidelines of the International Conference on Harmonization. A waiver for approval from the Spanish Medicines Agency was obtained, due to the classification of the fresh frozen plasma as a blood product. Full trial protocol is provided in the Supplementary Appendix.

### Procedures and Outcomes

Patients were registered, after giving informed consent to participate, in a web-based eCRF (ORACLE clinical), their baseline clinical data collected and then randomly assigned 1:1 to the investigational treatment, stratified by study site. CP had to be administered immediately after randomization (day 1). Inpatients were assessed daily until hospital discharge and at days 15 and 29. Assessments in discharged patients were performed either as outpatient consultations or by phone. The patient’s clinical status was recorded using the seven-category ordinal COVID-19 scale: 1, not hospitalized, no limitations on activities; 2, not hospitalized, limitation on activities; 3, hospitalized, not requiring supplemental oxygen; 4, hospitalized, requiring supplemental oxygen; 5, hospitalized, on non-invasive ventilation or high flow oxygen devices; 6, hospitalized, on invasive mechanical ventilation or ECMO and 7, death. The primary outcome was the proportion of patients in categories 5, 6 or 7 at day 15 of the study. Key secondary outcomes were time to improvement of one category on the ordinal scale; mean change in the ordinal scale from baseline to days 3, 5, 8, 11, 15 and 29; proportion of patients in categories 5, 6 or 7 at day 29; mortality at days 15 and 29; duration of hospital stay; number of days alive and free from oxygen support; number of days alive and free from mechanical ventilation. Serial naso/oropharyngeal swabs and blood samples were collected at days 3, 5, 8, 11, 15 and 29, and tested for SARS-CoV-2 RNA by RT-PCR assay. Serum samples were tested at the same time points for anti SARS-CoV-2 IgG. Neutralizing antibody titers were determined at baseline in those patients with positive IgG results. Serious adverse events (AE), grade 3 or 4 AE and infusion related AE (within 24 hours after administration) were collected. Investigators were instructed to actively monitor for the appearance of predefined AE of Special Interest: TRALI and ADE (antibody-dependent enhancement of infection).

### Statistical Analysis

A sample of 278 patients (139 per arm) was planned assuming 20% rate of worsening in the control group and an absolute reduction of 10% in the CP group, with 80% statistical power and 2.5% one-sided alpha level (5% two-sided). At the time of trial design there was substantial uncertainty about the expected proportion of patients worsening to categories 5, 6 or 7 at day 15. Therefore, a sample size re-estimation (at 60% of the trial size) and a series of futility and efficacy interim analyses were planned (at 20%, 40%, 60 and 80% of the trial size) using statistical boundaries based on rho family spending functions (with rho=7). The full Statistical Analysis Plan is available in the Supplementary Appendix.

The primary endpoint was planned to be estimated using a log-binomial regression model including center as a covariate, and predefining a contingency plan by using a Poisson regression model with log-link and robust variance estimator in case that the log-binomial would not fit. Due to the early stopping of the trial with a lower sample size and center distribution than expected, a Fisher test was used as a sensitivity analysis. For secondary outcomes, binary efficacy and safety outcomes are analyzed as described for the primary endpoint. The ordinal scale endpoint has been analyzed using the Wilcoxon rank-sum test (Mann–Whitney test) and a shift analysis using a logistic proportional odds model. Survival and median time to event [95% confidence interval - 95%CI-] were estimated by the Kaplan-Meier method and group comparisons done by log-rank test and the hazard ratios -HR- (95%CI) by the Cox model.

The trial was halted prematurely on July 10, 2020, shortly after the first interim analysis (cut-off date June 2, 2020), primarily driven by a drastic fall in recruitment, and despite having not reach futility or efficacy stop criteria.

## RESULTS

### Patients

Of the 87 patients included in the eCRF until July 10th, 81 underwent randomization, 38 were assigned to CP and 43 to the SOC group. Of the patients randomized to receive plasma, 37 (97.4%) received treatment as assigned (Figure 1). Patients characteristics are described in Table 1 and Table S1. Median age was 59 years and 54.3% of the patients were men. Median time interval between symptom onset and randomization was 8 days. At baseline 40 out of 81 patients (49.4%) tested positive for anti-SARS-CoV-2 IgG antibodies. There were no relevant between-group differences at baseline in demographic characteristics, baseline laboratory test results, distribution of ordinal scale scores, or concomitant treatments.

**Figure 1.**
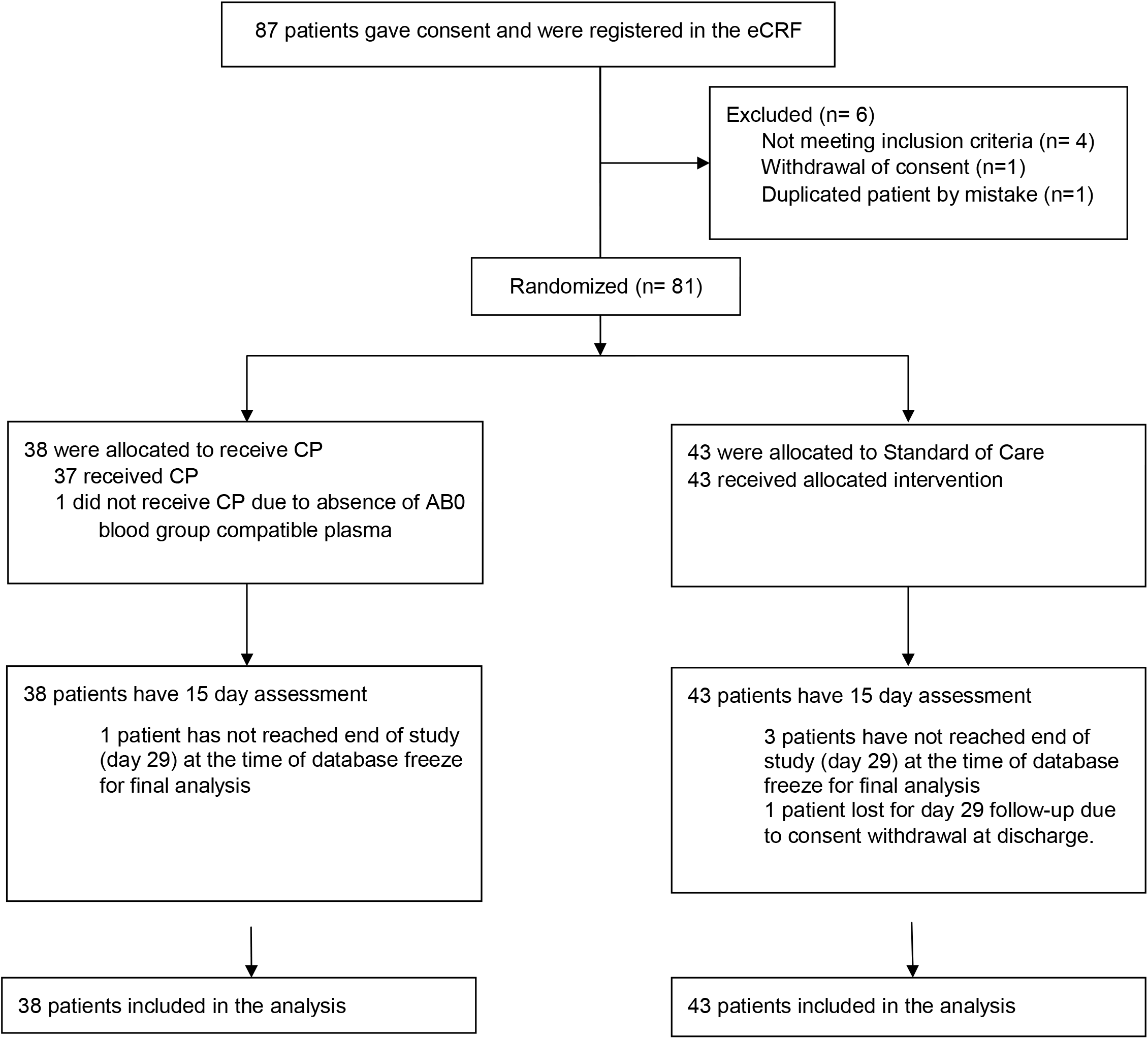
Enrollment and randomization

### Selection of Donors and Convalescent Plasma Characteristics

Out of 351 donors eligible for screening, 180 successful CP apheresis were performed after SARS-CoV-2 IgG positive results were obtained. Patients received CP units from 26 different donors. All administered units had neutralizing antibodies (VMNT-ID50: all titers >1:80, median titer 1:292, IQR 238-451; pseudovirus neutralizing ID50 assay: median titer 1:327, IQR 168-882, two CP units had ID50 titer <1:80). Supplementary information in Figure S1 and Tables S2 and S3.

### Primary Outcome

Patients assigned to CP had a lower rate of worsening at 15 days than patients receiving SOC only. Progression to categories 5-7 was 0 out of 38 patients (0%) in the CP group versus 6 out of 43 patients (14.0%) in the control group (Table 2; p=0.57, not statistically significant according to the originally planned analysis; p=0.03 using Fisher test as a post-hoc sensitivity analysis given small numbers and the by center heterogenous distribution).

### Key secondary outcomes

Progression to categories 5-7 at day 29 appears also lower in the CP group (0 out of 38; 0%) than in the control group (7 out of 43 patients, 16.3%; Table 2). Mortality rates were 0% in the CP arm and 9.3% in the SOC group (4 out of 43 patients) at days 15 and 29. No statistical difference between groups is shown in overall survival (p=0.06), in time to first clinical deterioration defined as a one-category worsening in any of the daily assessments (p=0.07), time to improvement in one-category or time to discharge (Figure 2). A visual description of changes in ordinal scale is included as Figure 3. Complete tables with changes in the ordinal scale, days free from oxygen and ventilatory support are included as supplementary tables. Planned subgroup analyses according to patient baseline characteristics (duration of disease, seroconversion) or titer of neutralizing antibodies in the administered plasma were not performed due to the low sample size. Serial RNA detection results of SARS-CoV-2 by RT-PCR from naso/oropharyngeal swabs were obtained at the expected timepoints in 79.7% of the cases in the plasma group and 66.54% in the control group. Six patients in each group had positive RNA results in blood samples at inclusion. Cumulative rates of RNA negativization and quantitative estimation of viral load decrease, measured by Ct values, did not show differences between groups either in naso/oro-pharyngeal swabs or blood samples, (table S8).

**Figure 2.**
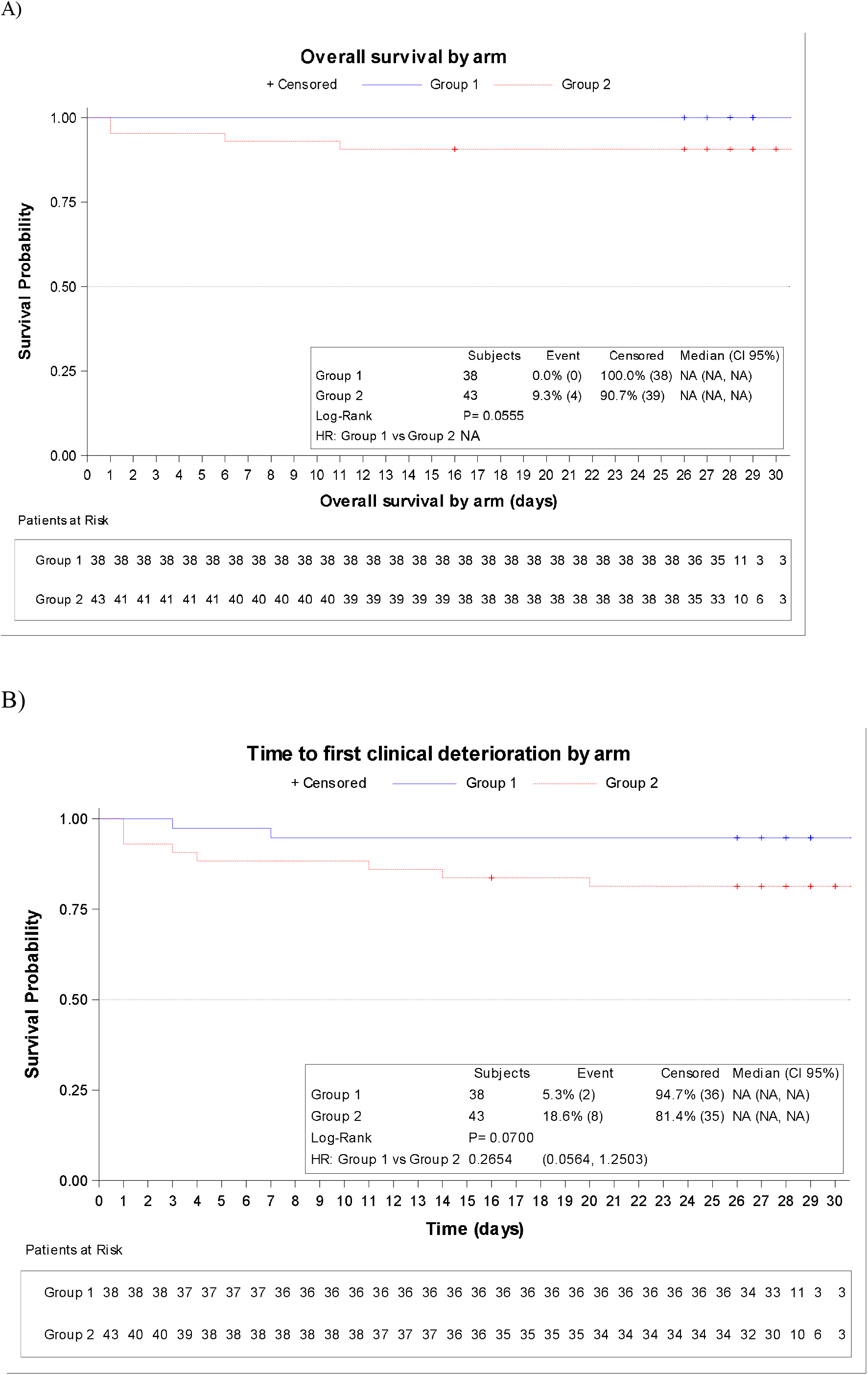
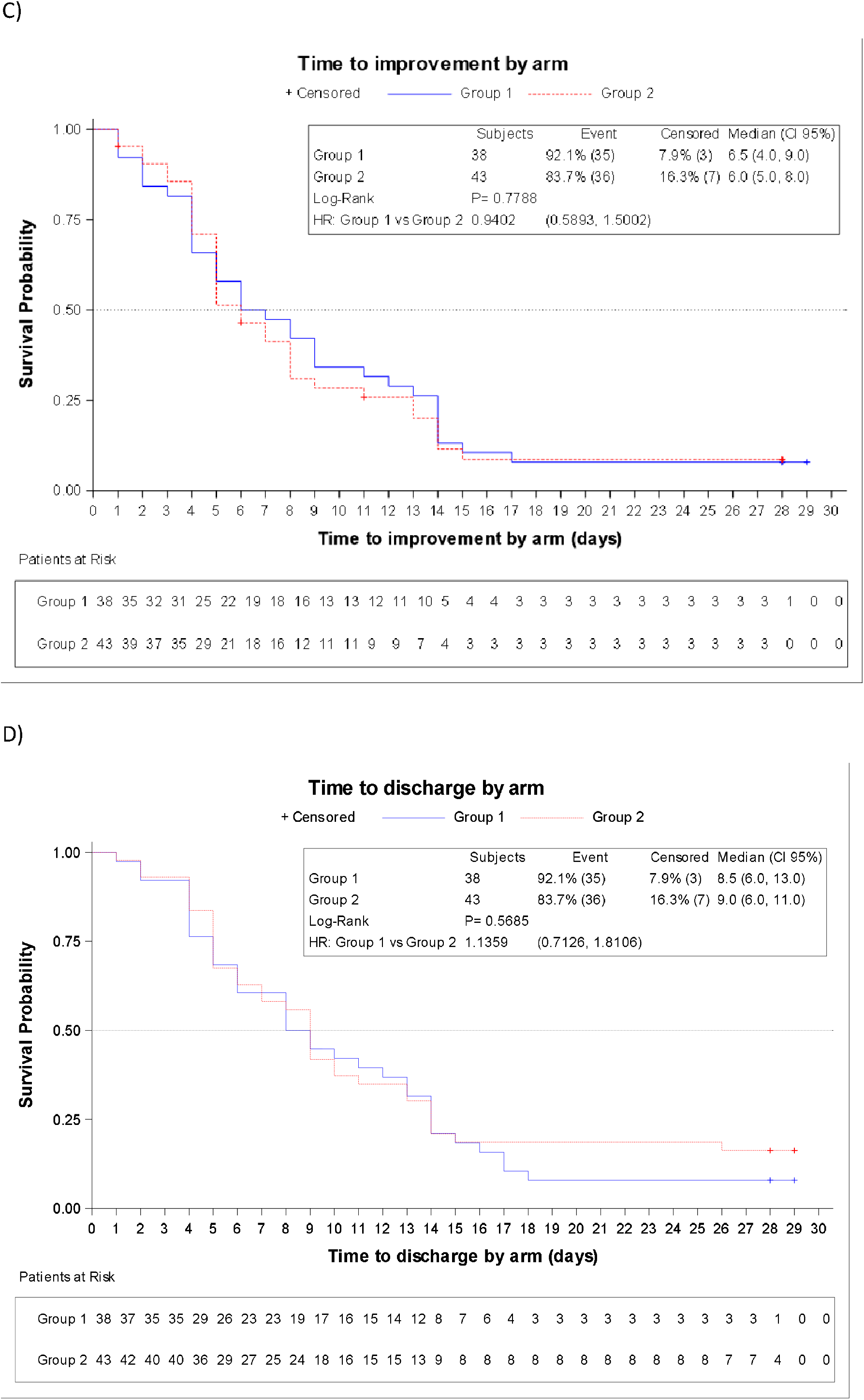
Kaplan-Meier estimates of secondary outcomes. A) Overall survival. Time to event curves for B) first clinical deterioration, C) first improvement of one category and D) Discharge.

**Figure 3.**
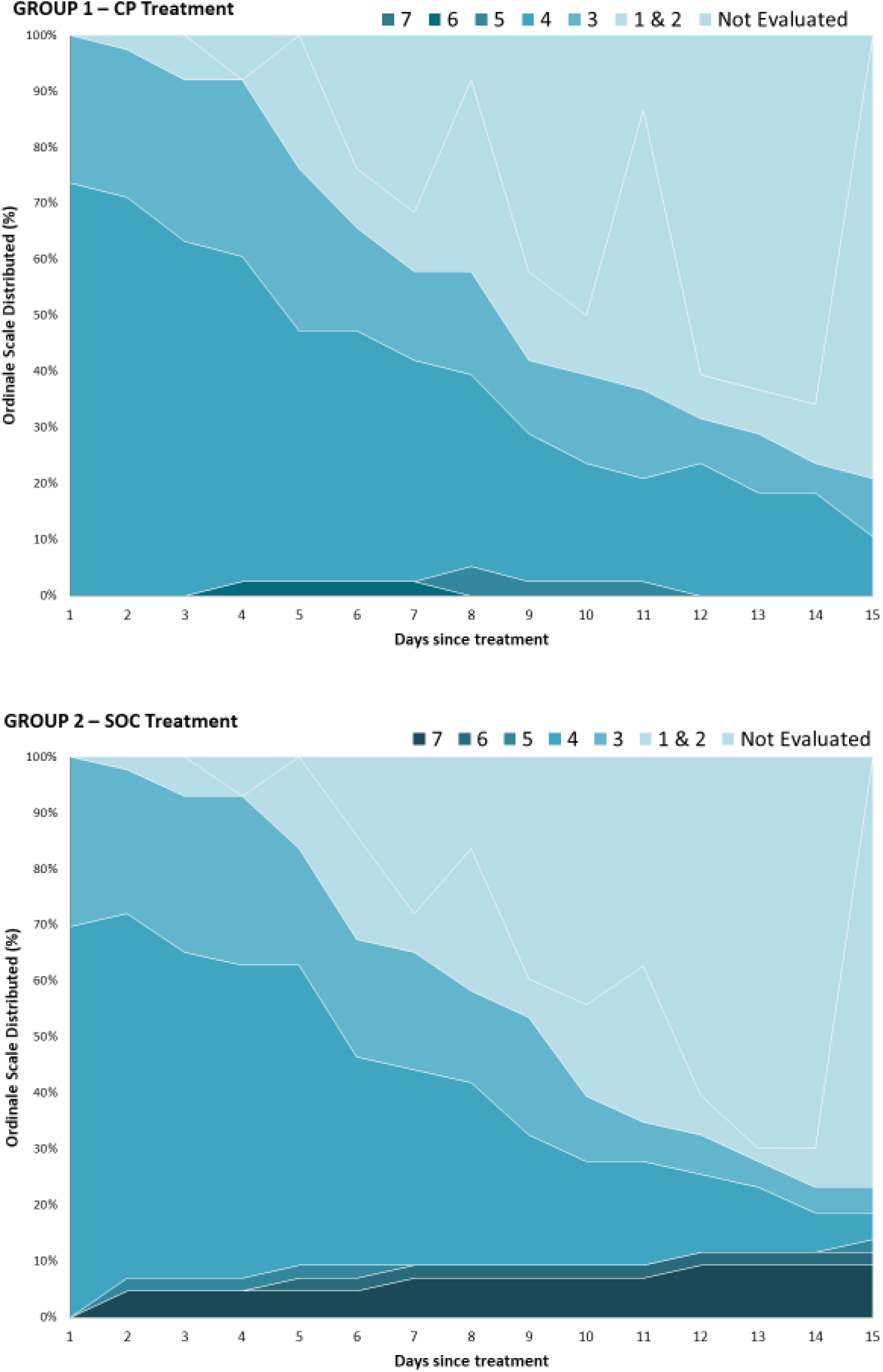
Distribution of the ordinal Scale results over time. All outpatients are shown on the same color: Not evaluated means outpatients not attending the hospital for a scheduled visit. More outpatients are evaluated on days 5, 8, 11 because of scheduled visits for samples collection. All outpatients were assessed on day 15 (main end-point). Outpatients assessed and categorized on levels 1 and 2 are combined.

### Safety

Sixteen serious or grade 3-4 AE were reported in 13 patients, 6 in the CP group and 7 in the SOC group (table S7). Two CP infusion-related AE and suspected TRALI were reported. In both cases, TRALI was ruled out after full assessment, including negative anti-HLA and anti-neutrophil antibodies in patients and donors. Both patients recovered without sequelae. None of the remaining events (n=14) were considered to be related to the CP. Five of the patients with a reported severe AE died due to their underlying disease (4 deaths within the study, 1 death after the end of the study, all in the SOC group). The remaining patients recovered without sequelae.

## DISCUSSION

Our study suggests that the addition of a single unit of CP to the SOC therapy of hospitalized COVID-19 patients may reduce the probability of disease progression to mechanical ventilation, ICU admission or death, both at day 15 (0% vs 14%) and at a longer follow-up on day 29 (0% vs 16.3%). No significant differences were found in other endpoints. These data complement two other RCT of CP in COVID-19, which were also halted prematurely. Li et al.^6^ saw no overall clinical benefit in 103 patients randomized at a median of 30 days from symptoms onset; in particular, no benefit in critically-ill patients admitted to ICU. However, the subgroup of patients with less-advanced clinical presentations had a better clinical improvement with CP than with SOC. A Dutch RCT by Gharbharan et al.^7^ was halted prematurely after 86 patients were enrolled. The vast majority of recruited patients (80%), including critically-ill patients in ICU, had SARS-CoV-2 IgG and neutralizing antibodies already at the time of randomization. At closure, the study showed no statistically significant differences between CP and SOC in mortality, clinical improvement at day-15 or other clinical outcomes. Unlike these RCT, ConPlas-19 excluded patients requiring high-flow oxygen devices or mechanical ventilation and targeted a population relatively earlier after symptom onset (median, 8 days; IQR 6-9). In spite of this, 49.4% of them still had antiSARS-CoV-2 IgG and neutralizing antibodies at baseline. Our design was driven by the strategic objective to prevent the larger group of less-severe hospitalized COVID-19 patients from requiring increased support and ICU admission at a time when the pandemic threatened to collapse our Health System. In addition, the scientific basis for a therapeutic effect of passive immunotherapy in COVID-19 and indirect evidence in other Severe Acute Respiratory Infections of Viral Etiology suggested that CP may be more effective when administered early after symptoms onset.^1,8^ Our data contribute to the evidence emerging from RCT, which despite the limitations of insufficient recruitment, bring together some shared conclusions valid for other potential studies and for clinical management. First, administration of CP in COVID-19 patients appears to be safe also in a well-monitored, randomized, controlled design. Second, they give support to studying CP earlier in the disease course in less-severe cases, rather than in advanced severe cases, in particular those in ICU.

This study has limitations. It is not blinded, but nevertheless, concealment of assignation was well preserved, all patients underwent a well-established standard treatment with no evident between-groups differences in treatment after enrolment. Its most relevant limitation is premature closure with a lower than expected number of patients and events. This clearly limits the identification and analysis of potential changes in many endpoints and patients’ subgroups and hampers our ability to draw definitive conclusions for the general population. With all three RCT available on CP in COVID-19 sharing low recruitment and premature closure, we must revise the unique difficulties that conduction of such RCT appear to face. Under the threat of a severe pandemic, the strong natural desire from clinicians, health systems and the society at large to prevent morbidity and mortality is commendable. However, in a situation of need, cognitive bias can make us all assume unproven efficacy without the rigor and evidence usually required.^9^ The role of the regulators in this context determines the course of events, in particular for an easily accessible treatment, such as CP. While regulatory authorities have made efforts during the pandemic to expedite the approval of RCT in COVID-19, they have been permissive about the use of CP in clinical practice (e.g. expanded access programs, observational studies, real-world experience).^10,11^ Tens of thousands of COVID-19 patients have been treated with CP based on the premise that RCT *“take significant time to produce results and will not be available for participation to all hospitals*”,^10^ despite the lack of evidence for efficacy, and the fact that such uncontrolled models lead to questionable estimates of treatment effects. The impact of these policies in Europe is hard to assess, as there have been no scientific reports from such observational studies since the EU program of CP in COVID-19 was released on April 4, 2020. Indirect evidence may suggest a negative impact, though. The Recovery trial reports a marginal number of patients randomized to CP despite several thousand being randomized to other therapeutic alternatives.^5^ In ConPlas-19, aside from the recent fall in recruitment related to pandemic control, out of 32 hospitals opened in 12 different Regional Health Departments in Spain, only 14 sites recruited any patients throughout the pandemic (supplementary appendix). In the US, an expanded access program has recently reported the first 35,322 COVID-19 patients treated with CP,^12^ suggesting apparent signatures of efficacy despite lacking untreated controls, and in a population of patients in which more than half were already in ICU at the time of treatment, which would be at odds with the lack of efficacy in this critically-ill setting in the few RCT available.^6,7^ While RCT in the US and worldwide accrue insufficient numbers, over 73,000 patients have been infused CP through open access in the US (www.uscovidplasma.org), and many more thousands are likely to have been treated in similar programs worldwide. If only a small percentage of these patients had been included in RCT, we would know by now whether CP is effective in COVID-19 at all, in which cases, and have robust evidence to make reliable recommendations to benefit future patients.^9^

In summary, CP appears to be safe in hospitalized COVID-19 patients who do not require high-flow oxygen devices or mechanical ventilation, and may reduce their probability of clinical deterioration, ICU admission or death. Formal recommendations for efficacy of treatment of COVID-19 patients with CP cannot be concluded from the evidence currently available including this RCT. Despite limitations, the authors decided to close and report the first phase of the trial as a better service to the scientific community. Our results shall be included in an ongoing initiative for pooling data from individual RCT of CP in COVID-19 (http://nyulmc.org/compile).^13^ We hope that this evidence will help the scientific community design and actively recruit in new revised RCT of CP in COVID-19 patients, likely in earlier forms of the disease, as the combined results from basic scientific knowledge and available RCT suggest that very severe COVID-19 cases, in particular those already in ICU and who are likely to have already developed neutralizing antibodies, do not benefit from CP. Generating robust evidence of the role of CP in these patients will be the best way, and perhaps the only way, to recognize the generosity of so many volunteer CP donors and to respond to the commendable strong desire of the society for reliable treatments for COVID-19 with definitive answers.

## Data Availability

All data will be available, including individual patient data after deidentification

## ACKNOWLEDGEMENTS

The authors would like to acknowledge the generous contributions of so many COVID-19 patients who either donated their plasma after recovering from the disease or agreed to participate as subjects in this study, as well as of so many healthcare professionals who, undeterred by the difficulties of the pandemic, helped looking after these patients and generating scientific evidence. We would like also to acknowledge the contribution of the members of the Data Safety Monitoring Board (Arantxa Sancho-Lopez, Emilio Ojeda, José Ríos, Juan Antonio Vargas and Carlos Vilches). This research is funded by the Government of Spain, Ministry of Science and Innovation, Instituto de Salud Carlos III, grant number COV20/00072 (Royal Decree-Law 8/2020, of 17 March, on urgent extraordinary measures to deal with the economic and social impact of COVID-19), co-financed by the European Regional Development Fund (FEDER) ‘‘A way to make Europe’’ and supported by SCReN (Spanish Clinical Research Network), ISCIII, project PT17/0017/0009. Clinical trial insurance coverage was kindly donated by MARCH RS Correduría de Seguros y Reaseguros. Mikel Mancheño-Losa holds a “Río Hortega” research contract (expte. CM19/00226).

## Notes

### Competing Interest Statement

The authors have declared no competing interest.

### Clinical Trial

NCT04345523

### Author Declarations

Research Ethics Committee of the Hospital Universitario Puerta de Hierro Majadahonda

### Summary of Updates

Abstract revised to delete a non significant p value for a secondary endpoint that had been left by mistake after removal of other p values from the abstract. Minor additional editing changes: Authors list in a paragraph instead of a list Supplementary Appendix revised to add some missing team members and correct fig S1, where a box was inadvertently missed

